# Exploring causal relationships and shared genomic signatures of coronary artery disease and stroke

**DOI:** 10.1101/2025.04.02.25325075

**Authors:** Tania Islam, Jian Zeng, Mohammad Ali Moni

## Abstract

Coronary artery disease (CAD) has been shown to be associated with stroke risk. However, the causal genetic overlap and shared genetic architecture underlying CAD and stroke remain unknown. Here, we aim to identify shared genetic relationships between CAD and stroke, focusing on causal genetic overlap, causal directions, and novel risk loci. Using MiXeR, we identified substantial causal variants (∼900) shared between CAD and stroke. LD score regression analysis showed a significant positive genetic correlation (rG = 0.49, P = 1.4 × 10^−48^) between CAD and stroke. We conducted Mendelian randomization analyses to estimate the potential causal association between CAD and stroke. We observed the significant causal association of CAD with the risk of stroke, independent from its common confounding factors, including type 2 diabetes, atrial fibrillation, BMI, smoking, and educational attainment. Multi-traits GWAS meta-analysis identified novel SNPs, while Summary-based Mendelian randomization analysis (SMR) was performed to identify causal risk genes linked to CAD and stroke. We identified 14 overlapping novel independent loci. SMR analysis further identified putative causal genes shared between CAD and stroke. This study advances our understanding of the shared genetic architecture and causal association between CAD and stroke risk, potentially indicating a common molecular mechanism underlying both conditions.

## Introduction

Stroke is a multifaceted neurological disorder that poses a serious threat to world health and ranks as the second leading cause of mortality. It accounts for around 12.2 million new cases annually, and 6.5 million people die due to stroke [1]. Coronary artery disease (CAD) is defined by the formation of atherosclerotic plaque in the inner walls of the coronary arteries, which accounts for approximately 7 million deaths [2]. The accumulation of plaques in the arteries blocks the circulation of oxygen-rich blood to vital organs, including the heart muscles and brain. Although stroke and CAD share some common risk factors [3], there are some distinct factors as well such as high cholesterol is more associated with CAD while hypertension is strongly linked to stroke [4–6]. Clinical and observational studies have shown stroke is more prevalent in patients with CAD as having CAD may exacerbate the atherosclerotic deposition in cerebral vascular systems or cardiogenic embolism [7, 8].

Both environmental and genetic factors also contribute to the development of CAD [9, 10] and stroke [11]. Family and twin studies have identified a heritability of 40% to 60% for CAD [10], and ∼40% heritability for stroke [12]. Past evidence also indicated the presence of shared heritability between CAD and stroke [13]. Recent large-scale genome-wide association studies (GWAS) have found > 200 risk loci associated with CAD [14] and >80 risk loci associated with stroke and stroke subtypes [15]; both CAD and stroke also shared some susceptible loci. Previously, genome-wide association and meta-analysis studies using METASTROKE (12,389 IS cases and 62,004 controls) and CARDIoGRAM (22,233 CAD cases and 64,762 controls) GWAS revealed several genetic variants for CAD that were also associated with ischemic stroke, indicating a similar genetic element enhance the risk of both CAD and stroke [16]. Despite much research, the causal associations of CAD on stroke risk are not completely known, as the observational associations are often hindered by confounding or reverse causations. In addition, the causal genetic overlap and shared genetic architecture of CAD and stroke remains unknown.

In this study, we have investigated the shared genetic architecture and causal association of CAD on stroke risk, leveraging large-scale GWAS datasets. We conducted conditional and joint analyses to account for the potential role of confounding factors, such as type 2 diabetes, in the observed associations between CAD and stroke. Cross-disorder GWAS meta-analysis was performed to identify novel risk loci associated with CAD and stroke, advancing our understanding of their common genetic basis.

## Methods

### Study Design

Firstly, we used MiXeR tool to estimate polygenicity and identification of overlapping genetic architecture between CAD and stroke. Next, we performed a linkage disequilibrium score regression (LDSC) study between CAD and stroke to evaluate the SNP-level genetic correlation. Secondly, we performed multi-trait conditional and joint analysis (mtCOJO) to avoid other risk factors that affects CAD and stroke. Then, we used Mendelian randomization (MR) analysis to ascertain bidirectional causal relationship between CAD and stroke. Third, we performed a muti-trait meta-analysis (MTAG) to boost the statistical power of individual GWAS datasets and facilitate the identification of novel risk loci associated with each trait. To identify novel independent lead SNPs shared between CAD and stroke, we performed functional mapping and annotation of the MTAG GWAS of T2D and stroke separately. Fourth, we performed summary data-based Mendelian randomization (SMR) analysis by integrating GWAS summary statistics and eQTL studies to identify putative functional causal genes and their role in CAD and stroke. Figure 5.1 illustrates the comprehensive research design of this study.

**Figure 1.**
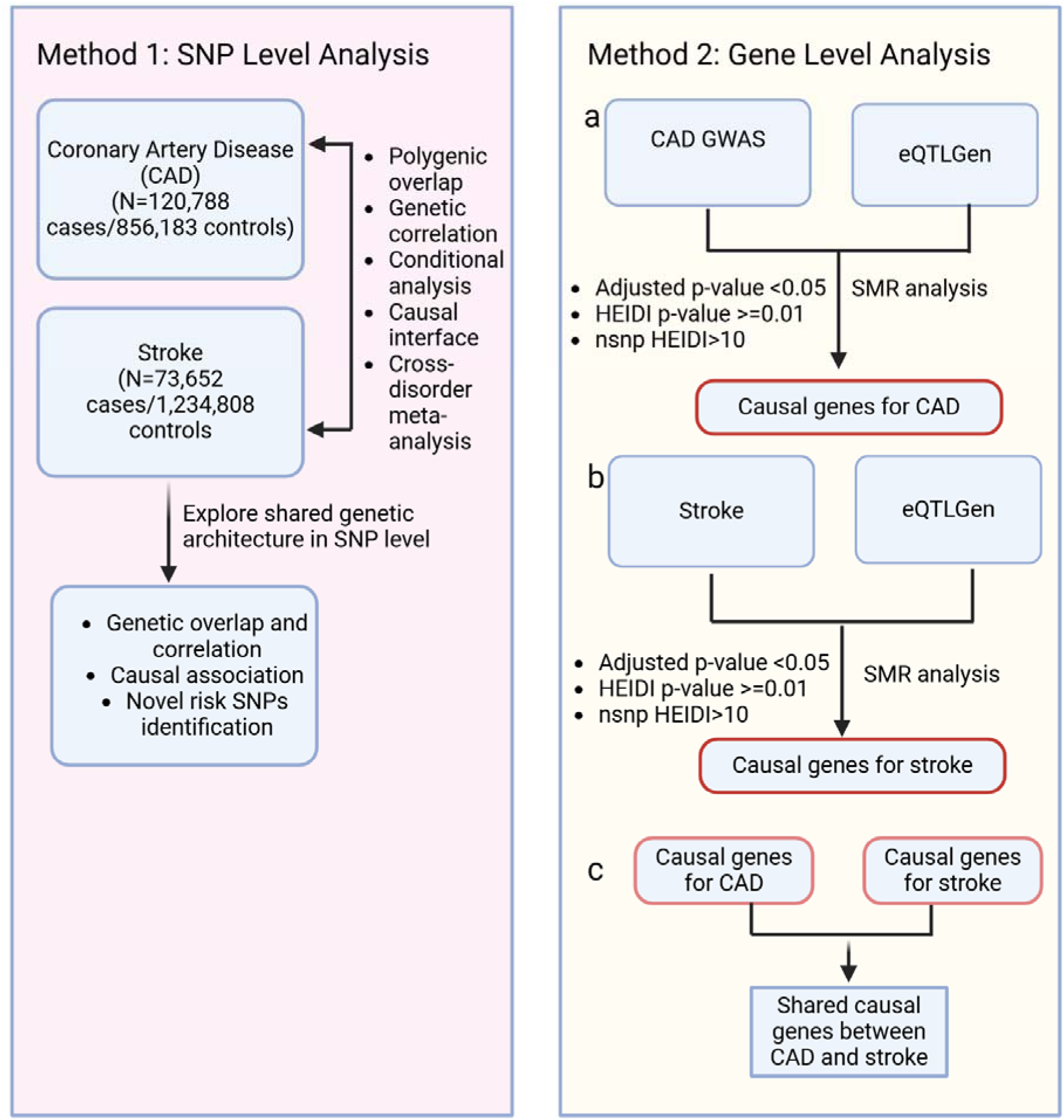
The schematic diagram represents the overall study design. In Method 1, we performed SNP-level analysis to identify genetic correlation, causal association, and novel independent risk loci shared by CAD and stroke. In Method 2, we conducted gene-based analysis to identify potential causal genes associated with CAD and stroke.

### Description of study samples

The GWAS summary statistics for CAD (GCST90132314) was obtained from the GWAS catalogue (https://www.ebi.ac.uk/gwas/), accessed on 15^th^ August 2023 which includes 120,788 European ancestry cases, and 856183 European ancestry controls [14]. Additionally, we got the GIGASTROKE consortium’s stroke GWAS summary data (GCST90104539) from the GWAS catalogue (https://www.ebi.ac.uk/gwas/) (accessed on 05^th^ April 2023), which includes 1,234,808 healthy controls of European ancestry and 73,652 stroke cases [15]. We performed a quality control analysis of these GWAS data to remove variants with MAF (<0.01), low imputation score (<0.60), out-of-range p-values, and mismatched allele pairings from the analysis.

### Quantification of shared genetic overlap between CAD and stroke

We applied MiXeR v1.3, a statistical tool of causal mixture model, to estimate the polygenicity, discoverability, and shared genetic overlap between CAD and stroke GWAS summary statistics. We performed two stages analysis: univariate analysis to characterize the genetic architecture of CAD and stroke individually to estimate the number of causal variants (polygenicity), their effect size variance (discoverability), and the total SNP heritability (h²_SNP), and bivariate analysis to quantify the polygenic overlap between CAD and stroke. Polygenic overlap was visualized using Venn diagrams, and statistical significance of the overlap was evaluated using a likelihood ratio test comparing the bivariate model to independent univariate models.

### Estimation of genetic heritability and global genetic correlation between CAD and stroke

To estimate the single trait liability-scale heritability (h2) and cross-trait genetic correlation (rg) between CAD and stroke, we used univariate and bivariate linkage disequilibrium score (LDSC) regression analyses, without intercept constrains. The LDSC model quantifies trait heritability and genetic correlation by regressing GWAS test statistics, such as Z-scores, onto the LD score for each SNP. This analysis used pre-computed LD scores from the 1000 Genomes European reference panel. Genetic correlation between CAD and stroke was considered significant if it exceeded the Benjamini–Hochberg false discovery rate (FDR) threshold of <0.05.

### A multi-trait conditional GWAS analysis to adjust the effect of covariates on CAD and stroke

To eradicate the effects of other covariates including type 2 diabetes, atrial fibrillation, body mass index, smoking, and education attainment on coronary artery disease and stroke, we performed multi-trait-based conditional and joint analysis (mtCOJO) [17, 18]. We used mtCOJO to condition GWAS summary data of CAD and stroke on the effect of AF, T2D, BMI, smoking, and EA respectively. After conditional analysis, we obtained trait-specific genetic variants which were used to evaluate the potential causality between CAD and stroke through Mendelian randomization analysis.

### Causal relationship assessment between CAD and stroke

To explore the putative causal relationship between CAD and stroke, we performed TwoSampleMR analysis (https://mrcieu.github.io/TwoSampleMR/)) to assess the possible causal impact of CAD on stroke. We performed MR analysis in three different conditions of exposure variables. First, to estimate the causal genetic relationship between exposure (CAD) and outcome (stroke), we performed MR analysis without conditioning the exposure (CAD) data. Second, we conditioned the exposure data (CAD) with type 2 diabetes to estimate the causal genetic relationship between CAD and stroke. Third, in this analysis, CAD was used as exposure variable-adjusted for atrial fibrillation (AF), type 2 diabetes (T2D), body mass index (BMI), smoking and education attainment (EA) while stroke served as the outcome. We did not condition the outcome (stroke) with T2D, AF, BMI, smoking and EA to avoid the collider bias.

We also performed reverse MR analysis, to estimate the causal effect of stroke on CAD risk. Here, first, we estimate the causal genetic relationship between exposure (stroke) and outcome (CAD), without conditioning the exposure variable. Second, we conditioned the exposure variable (stroke) with T2D and performed MR analysis to assess the causal genetic effects of stroke on CAD. Finally, the exposure data (stroke) was adjusted for type 2 diabetes (T2D), atrial fibrillation (AF), body mass index (BMI), smoking and education attainment (EA) while CAD served as the outcome.

For both MR analysis, we considered independent SPNs (LD clumping at *r*^2^ < 0.001) with genome-wide significant levels (p<5×10^−8^) as instrumental variables (IVs). To address and adjust for horizontal pleiotropy, we employed the Mendelian Randomization Pleiotropy Residual Sum and Outlier (MR-PRESSO) test. We also consider the SNP F-statistics value > 20 to quantifies the instrument variables (IVs) strength—higher values indicate strong instruments, while low values suggest potential weak instrument. We used inverse variance-weighted (IVW) method [19] as a main MR method to estimate the causal association. Additionally, for sensitivity analysis, we used weighted median [20], MR-Egger [21], and MR-PRESSO method [22] to determine the robustness of the result.

### MTAG analysis

To investigate the shared genetic basis of coronary artery disease (CAD) and stroke, we applied multi-trait analysis of genome-wide association studies (MTAG). This method generalizes inverse-variance-weighted meta-analysis by leveraging summary statistics from single-trait GWASs to derive trait-specific association [23, 24]. MTAG technique that considers sample overlap, imprecise genetic correlation, and genetic variability across traits with standard genetic bases [24]. We used MTAG version 1.0.8, CAD and stroke GWAS summary data were used as part of quality control (QC); MTAG excluded SNPs with absent values, minor allele frequency (MAF<0.01), out-of-range p-values, or mismatched allele pairings. After applying filtering criteria, MTAG retained 15,044,225 SNPs for coronary artery disease and 7,431,364 SNPs for stroke. After consolidating and eliminating strand-ambiguous SNPs, we obtained 6,079,007 SNPs for MTAG analysis. Following data consolidation and the removal of strand-ambiguous SNPs, 6,079,007 SNPs remained for MTAG analysis. We then performed a meta-analysis using the consolidated dataset, with Z-scores as the primary input for each trait. As part of the analysis, we estimated the genetic covariance matrix (Sigma) and precision matrix (Omega) to characterize the genetic association between CAD and stroke. The results revealed a genetic correlation of 0.487, highlighting substantial shared genetic architecture between the two traits. Quality control metrics showed that the GWAS mean chi-squared statistics were 1.602 for CAD and 1.112 for stroke, while the MTAG-adjusted chi-squared values were slightly elevated at 1.616 for CAD and 1.201 for stroke. The final MTAG results, including SNP effect sizes, standard errors, and significance values, were subsequently utilized for downstream analyses.

### Genomic loci characterization and independent SNP identification

We utilized the functional mapping and annotation (FUMA) platform (https://fuma.ctglab.nl) to identify the genomic loci and independent lead SNPs shared between CAD and stroke. For FUMA analysis, we separately input the MTAG-derived GWAS results for CAD and stroke. FUMA applied linkage disequilibrium (LD) clumping to identify independent SNPs (*r*^2^ < 0.1) and independent lead SNPs with no LD (*r*^2^ < 0.05) with a maximum p-value threshold of 5 x 10^-8^ [25]. The LD information was estimated using the 1000 Genomes Phase 3 (EUR) reference panel [25]. Next, to identify novel independent lead SNPs, we cross-referenced the GWAS Catalog to determine whether these SNPs had been previously reported as associated with CAD or stroke.

### Causal gene identification in CAD and stroke: Insights from summary data-based Mendelian randomization

To uncover the potential causal genes, whose expression is associated with CAD and stroke, we conducted a summary data-based Mendelian randomization (SMR) analysis. We performed SMR by integrating cis-expression quantitative trait locus (eQTL) data for whole blood from eQTLGen with the GWAS summary statistics [26]. The SMR approach ranks genes by leveraging the relationships between SNP effects on both gene expression and disease traits. Given that most genetic variants identified in GWAS lie in non-coding regions, it’s reasonable to infer that their influence on disease likely occurs primarily through the regulation of gene expression [13][27]. We used SMR to investigate how gene expression relates to both stroke and CAD. Additionally, we applied the heterogeneity in dependent instruments (HEIDI) test to distinguish between linkage and pleiotropy in the observed associations. To identify causal genes, we considered Bonferroni *P* value < 3 × 10^−6^ and *p*-value of HEIDI (*P*_HEIDI_ *>* 0.01) and minimum SNPs >10.

## Results

### Global genetic correlation and genetic overlap

We calculated heritability and global genetic correlations between CAD and stroke using LDSC. The SNP-based heritability for CAD and stroke at the liability scale was observed to be 0.096 (standard error, SE = 0.006) and 0.018 (SE = 0.001), respectively (Table 5.1). A strong global genetic correlation (rG = 0.496, P = 1.4 × 10−48) was found between CAD and stroke (Table 5.1).

**Table 1.** Estimation of heritability and genetic correlation of coronary artery disease and stroke.

SNP-based liability scale genetic heritability estimated from LDSC regression.
| Traits | Liability scale<br>h<sup>2</sup> | Lambda GC | Mean Chi<sup>2</sup> | Intercept |
| --- | --- | --- | --- | --- |
| Coronary<br>artery disease | 0.0968<br>(0.0066) | 1.4069 | 1.74 | 1.0127<br>(0.0194) |
| Stroke | 0.0184<br>(0.0015) | 1.1843 | 1.22 | 1.0702<br>(0.0068) |

SNP-based genetic correlation between CAD and stroke is estimated from LDSC regression.
| Trait1 | Trait 2 | rG (se) | P-value | Z-score | gcov_int_<br>se |
| --- | --- | --- | --- | --- | --- |
| Coronary<br>artery<br>disease | Stroke | 0.496<br>(0.033) | $1.404 \times 10^{-48}$ | 14.64 | 0.0062 |

Univariate MiXeR analysis revealed substantial polygenicity for both CAD and stroke, with CAD exhibiting greater polygenicity than stroke. The estimated number of trait-influencing variants for stroke was 1,000, while the number of trait-influencing variants for CAD was estimated at approximately 1,800. Next, we conducted bivariate MiXeR analysis and identified significant causal genetic overlap (Figure 5.2, Venn diagram), with 900 causal variants shared between CAD and stroke. The observed genetic overlap was also supported by strong genetic correlations (MiXeR r_g_ = 0.54). Condition Q-Q plot analysis showed the strong enrichment of shared variants in both directions, especially a steep leftward shift for most significant SNPs, suggesting shared genetic architecture between CAD and stroke (Figure 5.2). The log-likelihood plot confirmed robust MiXeR model fit (Figure 5.2).

**Figure 2.**
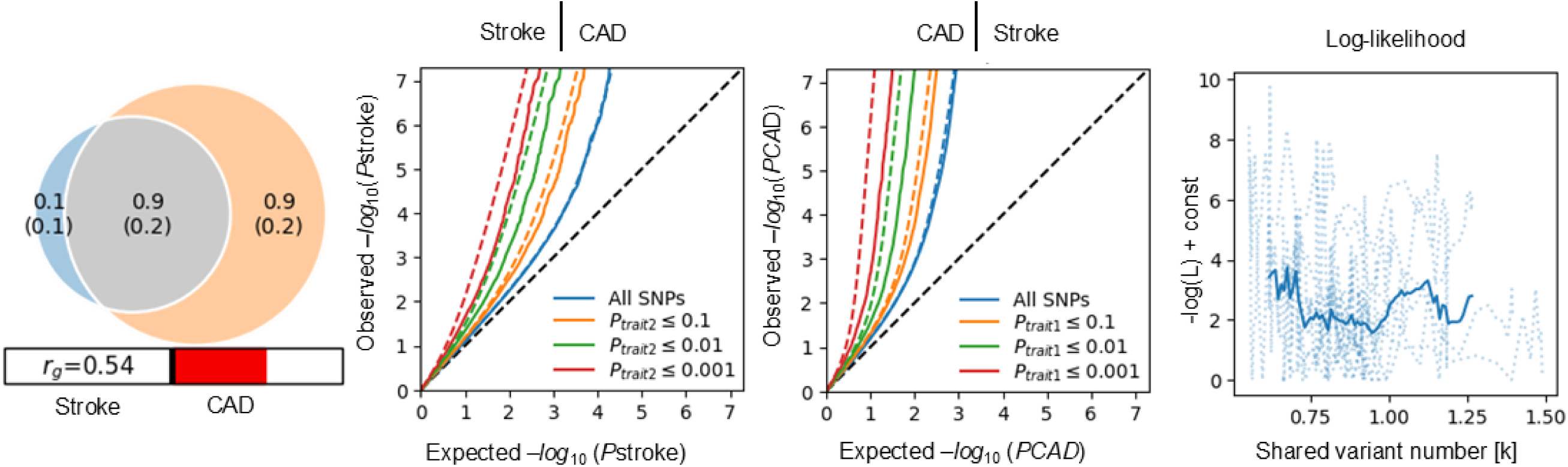
Venn diagram illustrates the polygenic overlap between stroke (blue) and CAD (orange). The numbers indicate the estimated proportion of causal SNPs (in percent), followed by standard errors in parentheses. The size of the circles reflects the degree of polygenicity. Conditional quantile-quantile (QQ) plot for stroke given CAD (stroke | CAD), showing observed vs. expected-log₁₀(P_stroke) values, with enrichment increasing as CAD association strength rises (all SNPs, P_CAD ≤ 0.1, ≤ 0.01, ≤ 0.001). Conditional QQ plot for CAD given stroke (CAD | stroke), displaying similar enrichment trends with less pronounced deviation. Log-likelihood plot as a function of shared variant number (in thousands [k]).

### Putative causal associations between CAD and stroke via Mendelian randomization

We performed Mendelian randomization analysis to assess the causal relationship between CAD and stroke. Initially, we selected 152 independent genome-wide significant SNPs (r2 <0.001) as instrumental variables associated with CAD (exposure) (sTable 5.1). Using the inverse variance method (IVW) as our primary approach, we identified a significant causal association between CAD and stroke, however, sensitivity analyses (via MR Egger) did not corroborate this finding (Table 5.2).

**Table 2.** Bidirectional causal genetic relationship between coronary artery disease and stroke using Mendelian randomization analysis without any adjustment.

| Exposure | Outcome | IVW |  | Weighted median |  | MR-Egger |  |  | MR-PRESSO |  |
| --- | --- | --- | --- | --- | --- | --- | --- | --- | --- | --- |
|  |  | OR | *P* | OR | *P* | OR | *P* | *P* Pleiotropy | OR | *P* |
| CAD | Stroke | 1.08 | $3.9 \times 10^{-10}$ | 1.07 | $1.12 \times 10^{-03}$ | 1.04 | 0.29 | 0.20 | 1.06 | $4.3 \times 10^{-06}$ |
| Stroke | CAD | 1.39 | $5.08 \times 10^{-28}$ | 1.35 | $1.57 \times 10^{-08}$ | 0.99 | 0.96 | 0.12 | 1.39 | $5.9 \times 10^{-06}$ |

Given that CAD is frequently confounded by cardiometabolic risk factors, particularly type 2 diabetes (T2D), we hypothesized that the genetic associations with CAD are biased by the confounding influence of T2D [28]. To address this, we utilized mtCOJO to adjust for the effects of T2D, thereby isolating the genetic contributions specific to CAD risk. Following this adjustment, we reran the MR analysis with 166 IVs (sTable 5.2) to assess the causal effect of CAD on stroke. Both the IVW and sensitivity analyses demonstrated a significant causal association between CAD and stroke risk (Table 5.3). We further conditioned CAD on atrial fibrillation, BMI, smoking, and educational attainment alongside T2D to eliminate residual confounding and pleiotropic effects, ensuring our IVs specifically capture the causal effect of CAD on stroke risk. This multi-trait adjustment, followed by MR with 155 IVs, consistently demonstrated a significant association (IVW OR = 1.19, P = 9.46 × 10⁻L¹), validated by sensitivity analyses (Table 5.4).

**Table 3.** Bidirectional causal genetic relationship between coronary artery disease and stroke using Mendelian randomization analysis with exposures adjusted for T2D.

| Exposure | Outcome | IVW |  | Weighted median |  | MR-Egger |  |  | MR-PRESSO |  |
| --- | --- | --- | --- | --- | --- | --- | --- | --- | --- | --- |
|  |  | OR | *P* | OR | *P* | OR | *P* | *P* Pleiotropy | OR | *P* |
| CAD (condition on T2D) | Stroke | 1.19 | $1.5 \times 10^{-41}$ | 1.18 | $7.6 \times 10^{-12}$ | 1.18 | $4.6 \times 10^{-06}$ | 0.94 | 1.18 | $1.6 \times 10^{-16}$ |
| Stroke (condition on T2D) | CAD | 1.37 | $5.6 \times 10^{-23}$ | 1.35 | $2.2 \times 10^{-08}$ | 1.08 | 0.71 | 0.28 | 1.37 | $2.8 \times 10^{-05}$ |

**Table 4.** Bidirectional causal genetic relationship between coronary artery disease and stroke using Mendelian randomization analysis, with exposures adjusted for T2D, AF, smoking, BMI, and EA.

| Exposure | Outcome | IVW |  | Weighted median |  | MR-Egger |  |  | MR-PRESSO (IVs:205) |  |
| --- | --- | --- | --- | --- | --- | --- | --- | --- | --- | --- |
|  |  | OR | *P* | OR | *P* | OR |  | *P* Pleiotropy | OR | *P* |
| CAD | Stroke | 1.18 | $9.46 \times 10^{-40}$ | 1.21 | $2.8 \times 10^{-15}$ | 1.22 | $2.03 \times 10^{-05}$ | 0.53 | 1.18 | $3.08 \times 10^{-15}$ |
| Stroke (9) | CAD | 1.19 | $3.2 \times 10^{-04}$ | 1.11 | $7.4 \times 10^{-02}$ | 0.43 | $1.9 \times 10^{-02}$ | 0.007 | 1.19 | $5.6 \times 10^{-02}$ |

To exclude the possibility of reverse causation—where stroke might drive CAD risk— we conducted bidirectional Mendelian randomization (MR) analysis, treating stroke as the exposure and CAD as the outcome. Using instrumental variables (IVs) robustly associated with stroke, we performed this reverse MR, but the results were inconclusive. Sensitivity analyses, including weighted median and MR-Egger, showed no consistent evidence of a significant causal effect of stroke on CAD (sTables 5.4–5.6, Tables 5.2–5.4). Multi-trait GWAS meta-analysis and causal genes

We conducted a multi-trait GWAS meta-analysis (MTAG) combining CAD and stroke GWAS datasets to enhance statistical power. After quality control, we analyzed 6,079,007 SNPs from up to 976,971 CAD and 1,308,460 stroke individuals. MTAG revealed a genetic correlation (rg) of 0.49 between CAD and stroke, suggesting shared genetic pathways account for ∼49% of risk overlap. Using MTAG outputs in FUMA, we identified 368 independent significant SNPs (r² < 0.1) for CAD (sTable 5.7) and 107 for stroke (sTable 5.8), with 47 common (sTable 5.9) and 14 novel (Table 5.5). Additionally, we found 280 lead SNPs (r² < 0.05) for CAD (sTable 5.10) and 94 for stroke (sTable 5.11), including 36 shared (sTable 5.12) and 9 novel SNPs per the GWAS catalog (Table 5.5).

**Table 5.**
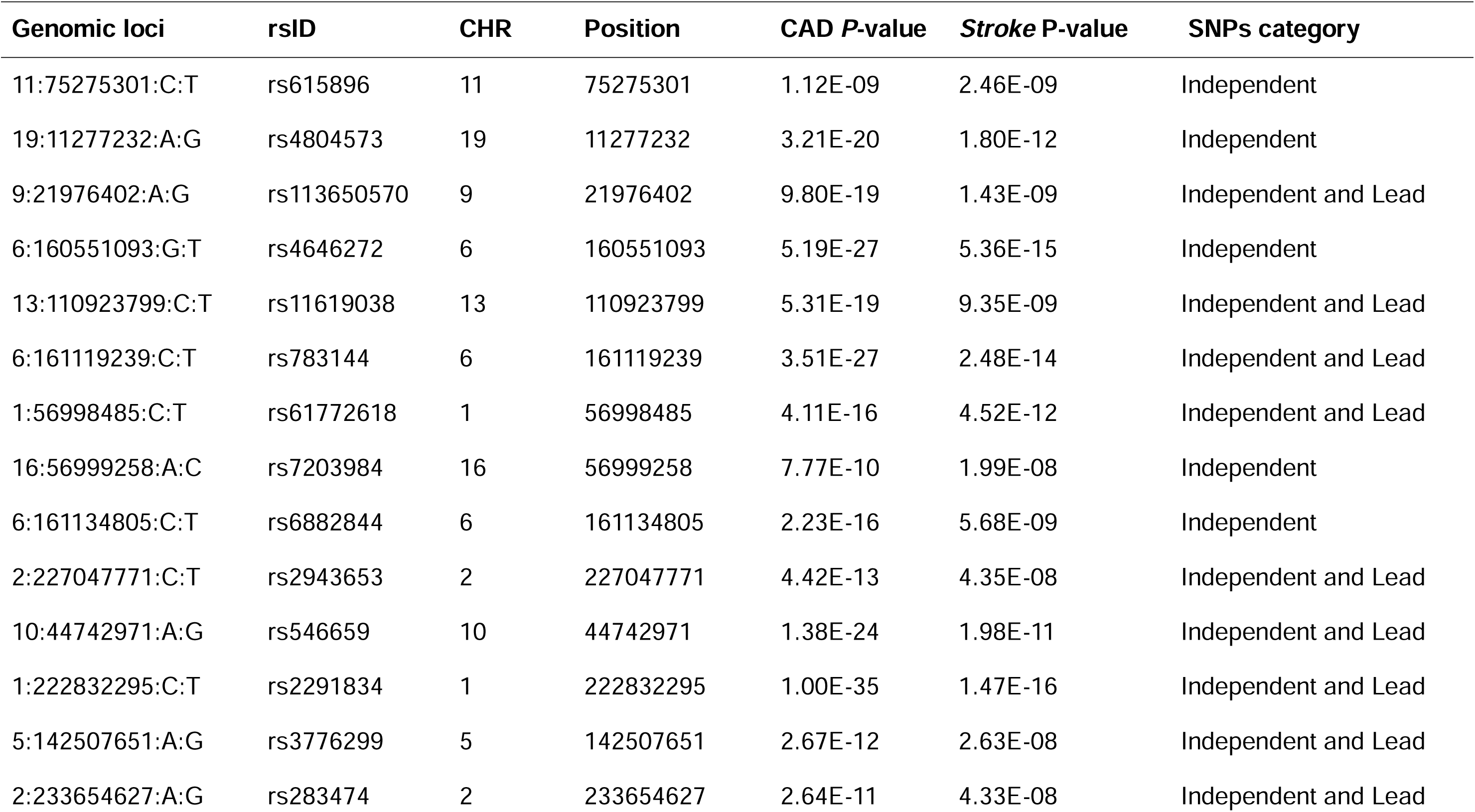
Common independent and lead SNPs associated with CAD and stroke.

| **Genomic loci** | **rsID** | **CHR** | **Position** | **CAD *P*-value** | **Stroke *P*-value** | **SNPs category** |
| --- | --- | --- | --- | --- | --- | --- |
| 11:75275301:C:T | rs615896 | 11 | 75275301 | 1.12E-09 | 2.46E-09 | Independent |
| 19:11277232:A:G | rs4804573 | 19 | 11277232 | 3.21E-20 | 1.80E-12 | Independent |
| 9:21976402:A:G | rs113650570 | 9 | 21976402 | 9.80E-19 | 1.43E-09 | Independent and Lead |
| 6:160551093:G:T | rs4646272 | 6 | 160551093 | 5.19E-27 | 5.36E-15 | Independent |
| 13:110923799:C:T | rs11619038 | 13 | 110923799 | 5.31E-19 | 9.35E-09 | Independent and Lead |
| 6:161119239:C:T | rs783144 | 6 | 161119239 | 3.51E-27 | 2.48E-14 | Independent and Lead |
| 1:56998485:C:T | rs61772618 | 1 | 56998485 | 4.11E-16 | 4.52E-12 | Independent and Lead |
| 16:56999258:A:C | rs7203984 | 16 | 56999258 | 7.77E-10 | 1.99E-08 | Independent |
| 6:161134805:C:T | rs6882844 | 6 | 161134805 | 2.23E-16 | 5.68E-09 | Independent |
| 2:227047771:C:T | rs2943653 | 2 | 227047771 | 4.42E-13 | 4.35E-08 | Independent and Lead |
| 10:44742971:A:G | rs546659 | 10 | 44742971 | 1.38E-24 | 1.98E-11 | Independent and Lead |
| 1:222832295:C:T | rs2291834 | 1 | 222832295 | 1.00E-35 | 1.47E-16 | Independent and Lead |
| 5:142507651:A:G | rs3776299 | 5 | 142507651 | 2.67E-12 | 2.63E-08 | Independent and Lead |
| 2:233654627:A:G | rs283474 | 2 | 233654627 | 2.64E-11 | 4.33E-08 | Independent and Lead |

We performed SMR analysis by integrating MTAG-generated GWAS summary statistics of CAD with eQTL summary data from the eQTLGen consortium, to identify potentially causal genes associated with CAD. We also performed SMR by integrating MTAG-generated GWAS summary statistics of stroke with eQTLGen data to identify causal genes associated with stroke. SMR and heterogeneity in dependent instruments (HEIDI) tests have identified 66 significant causal genes for CAD and 22 significant causal genes for stroke with Bonferroni corrected *P*-value< 3 × 10^-06^ (sTable 5.13 and sTable 5.14), respectively. Among the significant caused genes, we found 12 genes were common, and five genes: *RASD1, FAM109A, ADAM1A, UTP11, C6orf106* were novel for CAD and stroke. (Table 5.6).

**Table 6.** Identification of shared causal genes between CAD and stroke.

| **Genes** | **CHR** | **topSNP** | **p_SMR_CAD** | **P_HEIDI_CAD** | **p_SMR_stroke** | **p_HEIDI_stroke** | **CAD** | **Stroke** |
| --- | --- | --- | --- | --- | --- | --- | --- | --- |
| *RASD1* | 17 | rs12449964 | $1.81 \times 10^{-08}$ | 0.148 | $6.37 \times 10^{-07}$ | 0.697 | Novel | Novel |
| *FAM109A* | 12 | rs11065884 | $4.60 \times 10^{-09}$ | 0.13 | $2.08 \times 10^{-09}$ | 0.056 | Novel | Novel |
| *ADAM1A* | 12 | rs12423572 | $2.86 \times 10^{-10}$ | 0.471 | $5.36 \times 10^{-07}$ | 0.152 | Novel | Novel |
| *UTP11* | 1 | rs61776719 | $7.95 \times 10^{-12}$ | 0.260 | $1.72 \times 10^{-07}$ | 0.165 | Novel | Novel |
| *C6orf106* | 6 | rs76967117 | $7.30 \times 10^{-11}$ | 0.577 | $9.91 \times 10^{-08}$ | 0.432 | Novel | Novel |
| *CWF19L2* | 11 | rs12224337 | $6.29 \times 10^{-08}$ | 0.075 | $2.15 \times 10^{-06}$ | 0.543 | Known | Novel |
| *SREBF1* | 17 | rs9891957 | $7.30 \times 10^{-11}$ | 0.686 | $4.19 \times 10^{-10}$ | 0.650 | Known | Novel |
| *ING1* | 13 | rs9521916 | $7.68 \times 10^{-08}$ | 0.12 | $1.09 \times 10^{-06}$ | 0.188 | Known | Novel |
| *LIPA* | 10 | rs1412444 | $8.06 \times 10^{-29}$ | 0.324 | $3.67 \times 10^{-16}$ | 0.398 | Known | Known |
| *CALCRL* | 2 | rs8176580 | $1.98 \times 10^{-06}$ | 0.085 | $1.41 \times 10^{-06}$ | 0.237 | Known | Known |
| *PEMT* | 17 | rs12947517 | $1.24 \times 10^{-08}$ | 0.025 | $2.25 \times 10^{-07}$ | 0.028 | Known | Known |
| *RNU6-510P* | 1 | rs61776719 | $1.01 \times 10^{-11}$ | 0.258 | $1.87 \times 10^{-07}$ | 0.063 | Known | Novel |

## Discussion

This study has investigated shared genetic architecture and causal relationship between CAD and stroke leveraging the largest GWAS datasets of CAD and stroke. We observed significant genetic correlations and a significant causal association between CAD and stroke which is consistent with the prior observational studies [29, 30], but not vice versa, likely due to limited stroke IVs. Despite the high polygenicity of CAD than stroke [31], conditional analysis was performed to capture genetic association specific to CAD and identified that CA has a significant causal effect on stroke. Global genetic correlation and causal genetic overlap analysis showed significant genetic factors are shared between CAD and stroke, suggesting shared genetic components may play an important role in the progression of CAD and stroke. Cross-trait meta-analysis followed by FUMA-based annotation has revealed 14 novel independent SNPs, which were not previously reported. The novel lead SNP rs615896 is located in the *SERPINH1* gene, which encodes a heat-shock protein 47 (Hsp47) and is associated with the collagen secretion pathway [32]. *SERPINH1* gene also plays a significant role in the inflammation process and increases the risk of thrombosis [33]. The novel lead SNP rs113650570 is located in *CDKN2A/B* which encodes cyclin-dependent kinase inhibitors that play crucial roles in regulating the cell cycle and are implicated in vascular smooth muscle cell proliferation and are associated with cardiovascular disease [34]. Another novel lead SNP rs11619038 located within *COL4A1* genes which encodes the α1 chain of type IV collagen, an essential element of the basement membrane that preserves the structural integrity of blood vessels. The COL4A1/COL4A2 locus on chromosome 13q34 is a region associated with coronary artery disease [35]. Mutations in the *COL4A1* gene are associated with small-vessel disease and are implicated in a range of cerebrovascular disorders, including stroke [36]. Thus, the identified novel risk SNPs advanced our understanding of shared genetic relationships underlying CAD and stroke.

We identified five novel functional genes associated with CAD and stroke via SMR and HEIDI analyses. Among these, *RASD1* encodes ras-related dexamethasone-induced 1 protein which is involved in signal transduction pathways, with high expression in pituitary cell type [37], brain, heart, kidney, liver, and pancreas [38]. The *RASD1* also has a regulating role in cellular function, including circadian rhythms, adipogenesis, and obesity [37, 39]. Our analysis found that the downregulation of the *RASD1* is potentially causally linked with CAD and stroke. These shared genes highlight potential pleiotropic mechanisms underlying CAD-stroke comorbidity and warrant investigation as candidate biomarkers. Clinically, these findings support the development of genetic screening tools to identify CAD patients at heightened stroke risk, enabling targeted interventions to mitigate progression and improve outcomes.

We would like to acknowledge some limitations in our study. First, the sample size of stroke GWAS was limited which may affect the statistical power of the GWAS. Additionally, stroke and CAD GWAS were obtained from European ancestry which limited generalizability to other ancestries. Second, we only focused on the genetic contribution of CAD and stroke, but other environmental factors may have a role in disease development. Stroke is a heterogeneous disease with different subtypes, but our study does not distinguish between them, which may obscure subtype-specific genetic effects. Third, we identified the causal genes, but the experimental validation of the causal genes was not performed to confirm the biological mechanism linking CAD and stroke. Further wet lab validation is necessary to elucidate their causal role in disease progression.

## Conclusions

This study’s findings advanced our understanding of the potential causal shared genetic mechanisms underlying CAD and stroke. Our study suggests a significant genetic overlap and correlations exist between CAD and stroke. The MR analysis provided a significant causal role of CAD on stroke risk. However, reverse causality was not observed in our study due to limited instrumental variables in exposure (stroke), which suggests reverse causality could be assessed in the future when the largest stroke GWAS becomes available. MTAG analysis, followed by functional annotation of GWAS summary data revealed 14 novel independent SNPs shared between CAD and stroke, which could be utilized to predict high-risk individuals with stroke risk. Additionally, functional mapping of genes using SMR analysis identified that pleiotropic and/or causal genes are shared by T2D and stroke, which could be used as a potential biomarkers signature after web lab validation. Overall, the utilization of state-of-the-art statistical genetic and bioinformatic approaches advances our knowledge about the shared genetic aetiology of CAD and stroke, which could facilitate the development of new therapeutic approaches and points of intervention for these co-occurring diseases.

## Data Availability Statement

All data here analyzed is publicly available. The GWAS data used in this study are available in the GWAS Catalog. Stroke (https://www.ebi.ac.uk/gwas/studies/GCST90104539), CAD (https://www.ebi.ac.uk/gwas/studies/GCST90132314), AF (https://www.ebi.ac.uk/gwas/studies/GCST90204201), T2D (https://www.diagram-consortium.org/), BMI (https://www.ebi.ac.uk/gwas/studies/GCST009004), EA (https://ssgac.s3.amazonaws.com/EA4_additive_excl_23andMe.txt.gz), smoking (https://www.ebi.ac.uk/gwas/studies/GCST90042706)

## Code Availability

The code used in this study is available upon request.

## Acknowledgments

We thank the GIGASTROKE, DIAGRAM project, and GWAS catalog for making GWAS summary statistics data publicly available. I would like to acknowledge the Research Training scholarship from The University of Queensland, Australia.

## Authors contribution

All authors contributed to the conception and design of the study. TI conducted all analyses, prepared the figures and tables, and wrote the manuscript. JZ and MAM supervised the project and critically reviewed the manuscript.

## Conflict of Interest

“Authors declare no conflict of interest”.

## Figure legends

## Notes

### Competing Interest Statement

The authors have declared no competing interest.

### Funding Statement

This study did not receive any funding

